# Venous Thromboembolism in Ambulatory Covid-19 patients: Clinical and Genetic Determinants

**DOI:** 10.1101/2022.03.22.22272748

**Authors:** JQ Xie, A Prats-Uribe, Q Feng, YH Wang, D Gill, R Paredes, D Prieto-Alhambra

## Abstract

**Background:** Substantial evidence suggests that severe Covid-19 leads to an increased risk of Venous Thromboembolism (VTE). We aimed to quantify the risk of VTE associated with ambulatory Covid-19, study the potential protective role of vaccination, and establish key clinical and genetic determinants of post-Covid VTE.

**Methods:** We analyzed a cohort of ambulatory Covid-19 patients from UK Biobank, and compared their 30-day VTE risk with propensity-score-matched non-infected participants. We fitted multivariable models to study the associations between age, sex, ethnicity, socio-economic status, obesity, vaccination status and inherited thrombophilia with post-Covid VTE.

**Results:** Overall, VTE risk was nearly 20-fold higher in Covid-19 vs matched non-infected participants (hazard ratio [HR] 19.49, 95% confidence interval [CI] 11.50 to 33.05). However, the risk was substantially attenuated amongst the vaccinated (HR: 2.79, 95% CI 0.82 to 9.54). Older age, male sex, and obesity were independently associated with higher risk, with adjusted HRs of 2.00 (1.61 to 2.47) per 10 years, 1.66 (1.28 to 2.15), and 1.85 (1.29 to 2.64), respectively. Further, inherited thrombophilia led to an HR 2.05, 95% CI 1.15 to 3.66.

**Conclusions:** Ambulatory Covid-19 was associated with a striking 20-fold increase in incident VTE, but no elevated risk after breakthrough infection in the fully vaccinated. Older age, male sex, and obesity were clinical determinants of Covid-19-related VTE. Additionally, inherited thrombophilia doubled risk further, comparable to the effect of 10-year ageing. These findings reinforce the need for vaccination, and call for targeted strategies to prevent VTE during outpatient care of Covid-19.

## Introduction

Numerous hospital-based studies and case series have demonstrated a high risk of venous thromboembolism (VTE) in severe Covid-19 patients. A recent meta-analysis reported a pooled VTE rate of 14.7% and 23.2% among those admitted to hospital and in intensive care units, respectively.^1^ Additionally, emerging randomized controlled trials have shown the benefit-risk of anticoagulation for Covid-19 patients at different stages of the diseases,^2^ and clinical societies have recommended initiating routine antithrombotic therapy during hospital admission.^3^

In contrast, fewer clinical interventions have been implemented to prevent VTE amongst ambulatory patients with SARS-CoV-2 infection, partially due to conflicting findings on the association between infection and VTE risk, and mixed evidence of benefit from oral anticoagulation.^4^ Given the ongoing global rollout of vaccines, relaxation of public health restrictions, and widespread highly transmissible Omicron variant, the absolute number of milder Covid-19 cases managed in ambulatory settings continues to rise worldwide.^5,6^ These collectively suggest that prophylaxis, including timing and dosing regimens, requires further refinement, particularly in the outpatient setting.^2,7–10^ Moreover,^11^ a lack of insight into the impact of clinical, socioeconomic, and genetic risk factors for infection-related VTE persists.

This study aimed to (1) quantify the magnitude of acute VTE risk attributable to SARS-CoV-2 infection identified in ambulatory settings; and (2) investigate the clinical and genetic determinants of VTE risk after SARS-CoV-2 infection.

## Methods

We included UK Biobank (UKBB) participants from England who were alive on 01 March 2020. All participants provided written informed consent at the UKBB cohort recruitment. This study received ethical approval from UKBB Ethics Advisory Committee (EAC) and was performed under the application of 65397.

### Data sources and study cohorts

We obtained data from UK Biobank comprising multiple linked sources, including baseline surveys conducted between 2006-2010, individual genetic data, primary care electronic medical records, hospital inpatient data from Hospital Episode Statistics, diagnostic Covid-19 test data from Public Health England and death records from the national death registry (Office of National Statistics).

We curated an infected cohort by enrolling individuals with positive polymerase-chain-reaction SARS-CoV-2 results confirmed between 1 March 2020 and 30 September 2021. Participants who were never tested or only had negative test results were classified into the non-infected cohort. The index date was the date of the first positive specimen sample for the infected cohort, and a random date during the same calendar period for the matched non-infected. Participants with historical VTE or who used antithrombotic drugs one year before index dates were. Additionally, we excluded those in the infected cohort who tested positive in hospital settings.

### Inherited thrombophilia

Information on genotyping and imputation procedures in UK Biobank has been detailed in previous studies.^12^ Briefly, genome-wide single nucleotide polymorphisms (SNPs) were genotyped using two closely related purpose-designed arrays (the UK BiLEVE Axiom array and UK Biobank Axiom array). We defined inherited thrombophilia carriers as having any of two risk SNP variants in Factor V Leiden (rs6025) or Prothrombin G20210A (rs1799963). We also defined a positive genetic control exposure by calculating a 297-SNPs polygenic risk score (PRS) for VTE that did not include these two variants^13^ (see **Supplementary Appendix** for details).

### Covariates

We pre-specified a list of covariates for adjustment, including demographics (age, sex, ethnicity), socio-economic status measured by the Index of Multiple Deprivation (a summary deprivation measurement used in England containing seven aspects in crime, education, employment, health, housing, income and living environment),^14^ body mass index (BMI), medications prescribed within one year before the index date, and comorbidities recorded in primary care records (**Table** 1). The number of hospital admissions in the past one year (proxy of healthcare utilization) and vaccination status (no, partial 1-dose, or full 2-dose vaccination) were also studied.

**Table 1:** Demographic and clinical characteristics of participants, stratified by the SARS-CoV-2 infection status.

|  | Un-matched participants |  |  | Matched participants |  |  |
| --- | --- | --- | --- | --- | --- | --- |
|  | Non-infected | Infected | SMD | Non-infected | Infected | SMD |
| Number | 318,754 | 18,847 |  | 85,151 | 18,818 |  |
| Age, mean (sd) | 67.85 (8.04) | 64.32 (8.03) | 0.44 | 65.05 (7.92) | 64.33 (8.03) | 0.09 |
| Sex, male (%) | 132412 (41.5) | 8247 (43.8) | 0.045 | 36714 (43.2) | 8235 (43.8) | 0.012 |
| Ethnicity White (%) | 298406 (93.6) | 16590 (88.0) | 0.195 | 76981 (90.6) | 16578 (88.1) | 0.079 |
| Index of Multiple Deprivation, mean (sd) | 17.03 (13.51) | 19.87 (14.68) | 0.201 | 18.85 (14.59) | 19.83 (14.65) | 0.067 |
| Body mass index, mean (sd) | 27.04 (4.61) | 27.65 (4.83) | 0.129 | 27.43 (4.86) | 27.64 (4.81) | 0.044 |
| **Vaccination status** |  |  |  |  |  |  |
| Not vaccinated | 202661 (63.6) | 11140 (59.1) | 0.092 | 48144 (56.6) | 11117 (59.1) | 0.05 |
| Partial-vaccinated | 37471 (11.8) | 924 (4.9) | 0.25 | 3785 (4.5) | 923 (4.9) | 0.021 |
| Fully-vaccinated | 78622 (24.7) | 6783 (36.0) | 0.248 | 33076 (38.9) | 6775 (36.0) | 0.06 |
| **Medications (%)** |  |  |  |  |  |  |
| **(Past one year)** |  |  |  |  |  |  |
| Lipid lowering drugs | 86937 (27.3) | 4634 (24.6) | 0.061 | 20924 (24.6) | 4626 (24.6) | 0.001 |
| RAS inhibitors | 57790 (18.1) | 3296 (17.5) | 0.017 | 14645 (17.2) | 3290 (17.5) | 0.007 |
| Other anti-hypertensives | 26224 (8.2) | 1369 (7.3) | 0.036 | 6119 (7.2) | 1368 (7.3) | 0.003 |
| Proton pump inhibitors | 78004 (24.5) | 5268 (28.0) | 0.079 | 22564 (26.5) | 5247 (27.9) | 0.03 |
| Diabetes medicines | 16096 (5.0) | 1112 (5.9) | 0.037 | 4629 (5.4) | 1105 (5.9) | 0.018 |
| Antidepressants | 43411 (13.6) | 3189 (16.9) | 0.092 | 13403 (15.8) | 3180 (16.9) | 0.031 |
| Systemic glucocorticoids | 15031 (4.7) | 994 (5.3) | 0.026 | 4203 (4.9) | 984 (5.2) | 0.013 |
| Immunosuppressants | 3561 (1.1) | 203 (1.1) | 0.004 | 914 (1.1) | 203 (1.1) | <0.001 |
| Antineoplastic agents | 182 (0.1) | 14 (0.1) | 0.007 | 51 (0.1) | 14 (0.1) | 0.006 |
| **Coexisting conditions (%)** |  |  |  |  |  |  |
| **(Past years)** |  |  |  |  |  |  |
| Cancer | 31098 (9.8) | 1476 (7.8) | 0.068 | 7050 (8.3) | 1475 (7.8) | 0.017 |
| Malignant cancer | 1102 (0.3) | 63 (0.3) | 0.002 | 283 (0.3) | 63 (0.3) | <0.001 |
| Diabetes (uncomplicated) | 23095 (7.2) | 1496 (7.9) | 0.026 | 6396 (7.5) | 1490 (7.9) | 0.015 |
| Diabetes (end-organ damage) | 6651 (2.1) | 415 (2.2) | 0.008 | 1775 (2.1) | 414 (2.2) | 0.008 |
| Congestive heart failure | 1629 (0.5) | 90 (0.5) | 0.005 | 410 (0.5) | 90 (0.5) | 0.001 |
| Myocardial infarction | 645 (0.2) | 35 (0.2) | 0.004 | 156 (0.2) | 35 (0.2) | 0.001 |
| Cerebrovascular disease | 2426 (0.8) | 138 (0.7) | 0.003 | 639 (0.8) | 136 (0.7) | 0.003 |
| Peripheral vascular disease | 1317 (0.4) | 49 (0.3) | 0.026 | 224 (0.3) | 49 (0.3) | 0.001 |
| Liver disease (mild) | 1493 (0.5) | 81 (0.4) | 0.006 | 397 (0.5) | 81 (0.4) | 0.005 |
| Liver disease (moderate to severe) | 595 (0.2) | 46 (0.2) | 0.012 | 184 (0.2) | 45 (0.2) | 0.005 |
| COPD | 49824 (15.6) | 3309 (17.6) | 0.052 | 14373 (16.9) | 3294 (17.5) | 0.016 |
| Chronic kidney disease | 12746 ( 4.0) | 691 ( 3.7) | 0.017 | 3152 ( 3.7) | 688 ( 3.7) | 0.003 |
| Peptic ulcer | 6240 ( 2.0) | 419 ( 2.2) | 0.019 | 1827 ( 2.1) | 418 ( 2.2) | 0.005 |
| Rheumatoid arthritis | 8310 ( 2.6) | 511 ( 2.7) | 0.006 | 2276 ( 2.7) | 509 ( 2.7) | 0.002 |
| Dementia | 1873 ( 0.6) | 206 ( 1.1) | 0.055 | 786 ( 0.9) | 202 ( 1.1) | 0.015 |
| Hemiplegia | 244 ( 0.1) | 15 ( 0.1) | 0.001 | 65 ( 0.1) | 15 ( 0.1) | 0.001 |
| AIDS | 302 ( 0.1) | 17 ( 0.1) | 0.001 | 69 ( 0.1) | 17 ( 0.1) | 0.003 |
| **Hospital admissions (numbers, past one year)** | **0.30 (1.51)** | **0.32 (1.96)** | **0.014** | **0.29 (1.39)** | **0.31 (1.61)** | **0.015** |

SMD: Standardised mean difference, COPD: Chronic obstructive pulmonary disease, AIDS: Acquired immune deficiency syndrome

### Outcomes

Incident VTE, comprising either deep vein thrombosis or pulmonary embolism, was identified using ICD-10 codes based on hospital records. Eligible participants were followed up for up to 30 days after the index date, given that VTE occurring after 30 days were much less likely to be due to a direct consequence of SARS-CoV-2 infection.

### Statistical analyses

We used propensity score (PS) matching to minimize confounding in the study of the association between SARS-CoV-2 infection and VTE. We fitted multivariable logistic regression models to estimate PS as the probability of infection based on the predefined covariates. We then matched infected with non-infected individuals with a ratio of 1:5 based on PS values with a caliper width of up to 0.2 standard deviations of the logit of the PS, with exact-matching on index dates.^15,16^ We assessed covariate balance between the cohorts before and after matching using absolute standardized mean differences (SMD) and specified SMD>0.1 as relevant imbalances.^17^ Cause-specific Cox survival models were applied to estimate hazard ratios (HRs) for VTE according to exposure, in which death was considered a competing risk.^18^

To study clinical determinants, we fitted multivariable Cox models of 30-day VTE in the ambulatory Covid-19 cohort, including age, sex, ethnicity, socio-economic status, obesity (BMI less than vs equal or more than 30 kg/m^2^) and vaccination status. For the analysis of inherited thrombophilia’ effect on post Covid-19 VTE, we adjusted Cox models for age, sex and genetic ancestry (quantified by the first three principal components), assuming that genetic variants were independent of baseline characteristics. We introduced a positive exposure control (PRS for VTE) and a negative outcome (arterial thromboembolism [ATE]) experiment to detect residual confounding and potential unresolved bias.^19^

A sensitivity analysis was performed repeating the modelling of clinical determinants with VTE in the non-infected cohort, where no association is expected between vaccination and Covid-19-unrelated VTE.

All statistical tests were 2-sided, where a *P* = 0.05 or a 95% CI that did not contain unity were considered statistically significant for the primary analyses. All analyses and data visualizations were conducted using R statistical software. Genetic data management and quality controls were performed using Plink 1.9.^20^

## Results

### Baseline characteristics

Out of 407,311 UKBB participants, 26,210 were infected with SARS-CoV-2 between 1 March 2020 and 30 September 2021. After applying exclusion criteria, 21,724/ 26,210 (83%) infected and 318,754/ 407,311 (78.23%) non-infected were eligible for analyses (**Supplementary Results 1**). For all infections, 2,877 (13 %) and 18,847 (87 %) tested in hospital and outpatient settings, respectively. Only the latter were included for subsequent analyses.

Baseline characteristics by infection status are summarized in **Table 1**. Overall, ambulatory Covid-19 participants were younger than the uninfected (mean age [sd] 64.32 [8.03] vs 67.85 [8.04]), more likely male (43.8% vs 41.5%), from non-White ethnicity (12.0 % vs 6.4%), and more socioeconomically deprived and obese. After PS matching, almost all ambulatory Covid-19 cases (98.7%) were successfully matched to at least one non-infected counterpart, and all covariates became balanced. In addition, index dates and calendar periods were accurately aligned between the cohorts, as depicted in **Supplementary Results 2**.

### Association of SARS-CoV-2 infection with incident VTE

Figure 1. depicts cumulative incidence of VTE according to infection status, showing an early separation of the matched cohorts, with continued divergence over time. A total of 73 and 17 VTE events were seen within 30 days amongst the ambulatory Covid-19 patients and matched uninfected, corresponding to incidence rates of 51.00 and 2.61 per 1,000 person-years, respectively. Survival analyses (**Table 2**) suggested that SARS-CoV-2 infection was associated with a striking increase in VTE risk: HR: 19.49, 95% CI 11.50 to 33.05. The observed risk was more pronounced in the unvaccinated patients (HR: 29.37, 95% CI 15.11 to 57.08) and substantially mitigated in the fully vaccinated (HR: 2.79, 95% CI 0.82 to 9.54).

**Figure 1:**
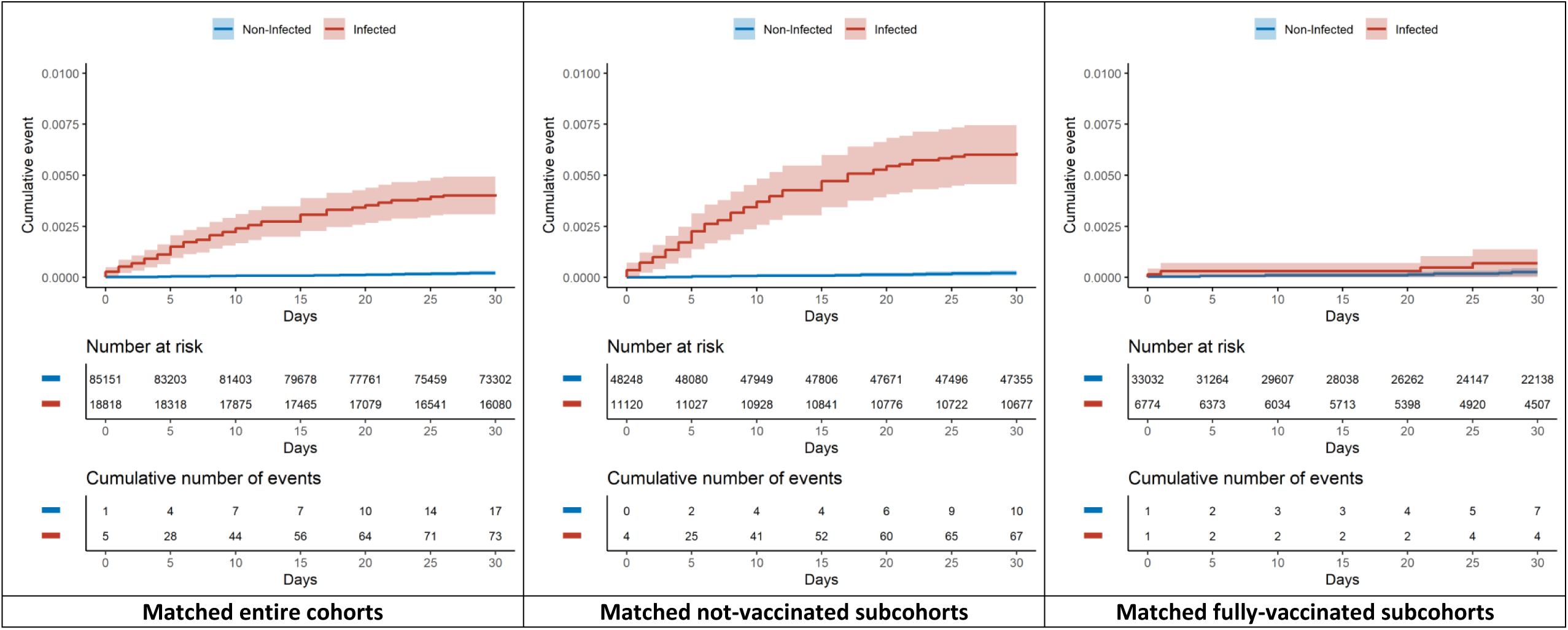
Cumulative incidence curves of venous thromboembolic events within 30 days.

**Table 2:** Associations between ambulatory Covid-19 and venous thromboembolic events, overall and stratified by vaccination status

|  | Matched Non-infected |  |  | Matched Infected |  |  | Hazard ratio<br>(95% CI) | P-value |
| --- | --- | --- | --- | --- | --- | --- | --- | --- |
|  | No. of<br>People | No. of<br>Events | Incidence<br>per 1000 PYs | No. of<br>People | No. of<br>Events | Incidence<br>per 1000 PYs |  |  |
| All participants | 85,151 | 17 | 2.61 | 18,818 | 73 | 51.00 | 19.49 (11.50 to 33.05) | <0.01 |
| **Subgroups\*** |  |  |  |  |  |  |  |  |
| Not vaccinated | 48,248 | 10 | 2.54 | 11,120 | 67 | 75.09 | 29.37 (15.11 to 57.08) | <0.01 |
| Fully vaccinated | 33,032 | 7 | 3.08 | 6,774 | 4 | 8.61 | 2.79 (0.82 to 9.54) | 0.10 |

\*No separate analysis was performed among the half-vaccinated (one does) subgroup due to the minimal sample size.

### Clinical determinants of post-Covid-19 VTE

The associations between socio-demographic and clinical factors (including vaccination status) and risk of post-Covid VTE are shown in **Figure 2**. Older participants had a higher risk, with an approximate doubling of risk per each 10-year increase in age (adjusted HR: 2.00, 95% CI 1.61 to 2.47). Men were at higher risk than women (adjusted HR: 1.66, 95% CI 1.28 to 2.15), and people with obesity at a higher risk than non-obese (adjusted HR: 1.85, 95% 1.29 to 2.64). These associations were equally seen for Covid-19-unrelated VTE in both direction and magnitude (**Figure 2; Supplementary Results 3**).

**Figure 2:**
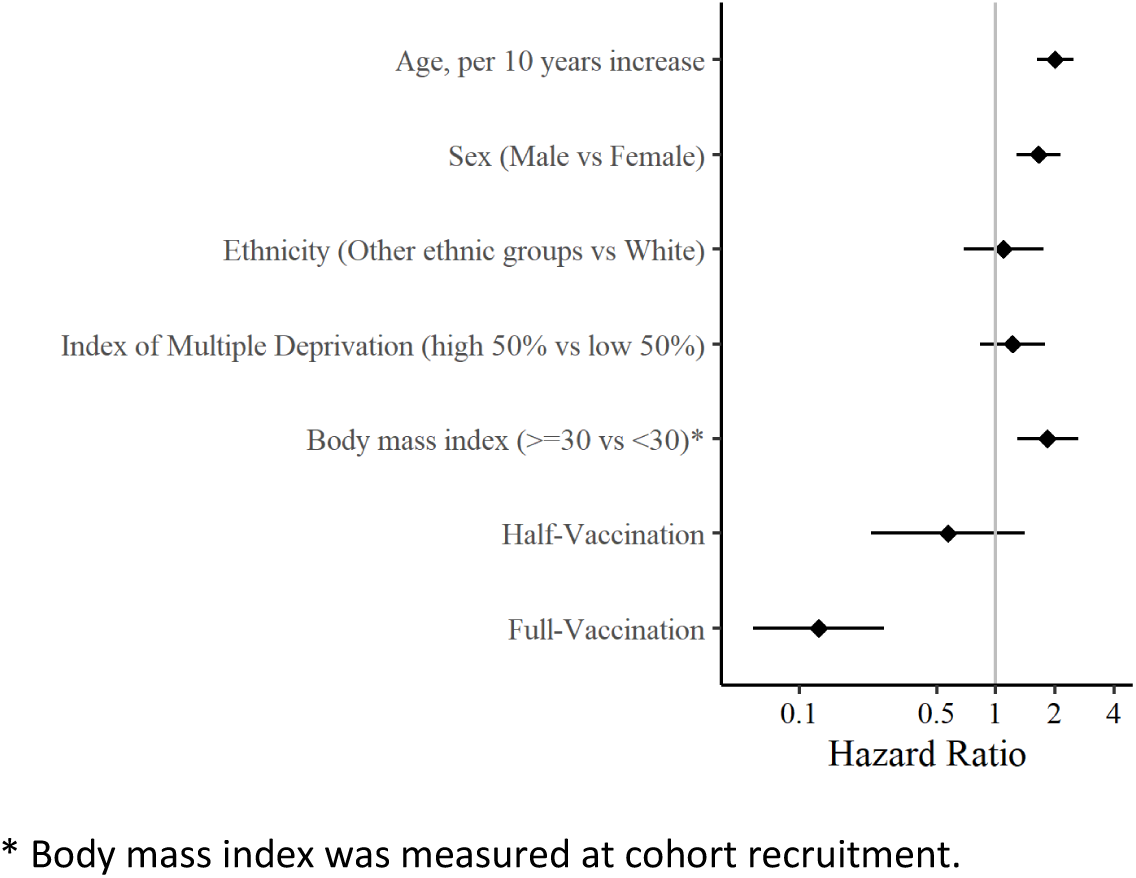
Associations of clinical factors and vaccination status with Covid-19-related VTE (Any VTE events in the ambulatory Covid-19 cohort). * Body mass index was measured at cohort recruitment.

Additionally, 2-dose vaccination was associated with a substantially lower risk of Covid-19-related VTE (adjusted HR: 0.13, 95% 0.06 to 0.28). A sensitivity analysis demonstrated no association between vaccination status and Covid-19-unrelated VTE (**Figure 2; Supplementary Results 3**).

### Inherited thrombophilia and post-Covid-19 VTE

Among 21,055 infected participants with complete genetic data, 1,287 (6.11%) had inherited thrombophilia, with 909 (4.32%) and 392 (1.86%) carrying risk variant/s of Factor V Leiden and Prothrombin G20210A, respectively (**Table 3**). The frequency of these genetic variations in the infected cohort was like that in the overall UKBB cohort and consistent with reports from previous literature (**Supplementary Results 4**). No differences in any of the measured covariates (except for ethnicity), including socio-demographics, medications or comorbidities, were observed when comparing those with vs without inherited thrombophilia (**Supplementary Results 5**).

**Table 3:** Effects of inherited thrombophilia on venous and arterial thromboembolism amongst all SARS-CoV-2 infected participants.

| Exposure | Frequency (%) | Primary outcome |  |  | Negative outcome |  |  |
| --- | --- | --- | --- | --- | --- | --- | --- |
|  |  | Venous thromboembolism |  |  | Arterial thromboembolism |  |  |
|  |  | Unadjusted HR.<br>(95% CI) | Adjusted HR<br>(95% CI) | *P*<br>value | Unadjusted HR<br>(95% CI) | Adjusted HR<br>(95% CI) | *P*<br>value |
| Inherited thrombophilia | 6.11 | 1.82 (1.02 to 3.23) | 2.05 (1.15 to 3.66) | 0.01* | 0.94 (0.51 to 1.73) | 0.98 (0.53 to 1.80) | 0.95 |
| Factor V Leiden | 4.32 | 1.97 (1.03 to 3.76) | 2.17 (1.13 to 4.15) | 0.02* | 0.97 (0.48 to 1.98) | 0.99 (0.49 to 2.01) | 0.97 |
| Prothrombin G20210A | 1.86 | 1.31 (0.42 to 4.11) | 1.52 (0.48 to 4.79) | 0.45 | 0.84 (0.27 to 2.62) | 0.91 (0.29 to 2.84) | 0.86 |
| **Positive control** |  |  |  |  |  |  |  |
| Continuous PRS<sup>§</sup> | NA | 1.29 (1.08 to 1.54) | 1.34 (1.11 to 1.60) | <0.01* | NC | NC | NC |
| Categorical PRS<sup>‡</sup> | 6% | 1.39 (0.73 to 2.66) | 1.54 (0.80 to 2.95) | 0.20 | NC | NC | NC |

HR: Hazard ratio, PRS: polygenic risk score, NA: Not available, NC: Not calculated
<sup>§</sup>Per 1-SD increase of PRS, <sup>‡</sup> top 6% of PRS vs the lower, \* statistical significance
To set an informative, positive exposure arm, we classified people as low and high polygenic risk groups according to their PRS value with a cut-off point at the 94% percentile. This threshold was chosen based on the proportion of inherited thrombophilia in the general UK Biobank population (~6%).

Participants with inherited thrombophilia had a higher risk of VTE following SARS-CoV-2 infection than those without (adjusted HR: 2.05, 95% CI 1.15 to 3.66). For each risk variant, the adjusted HR was 2.17 (95% CI 1.13 to 4.15) for Factor V Leiden carriers and 1.52 (95% CI 0.48 to 4.79) for Prothrombin G20210A carriers. Also, individuals with higher PRS values had greater VTE risk: adjusted HR per 1-SD increase of PRS 1.33, 95% 1.11 to 1.59 (**Table 3**). As expected, no associations were observed between inherited thrombophilia and the negative control outcome ATE, with adjusted HR ranging from 0.91 (95% CI 0.29 to 2.84) to 0.99 (95% CI 0.49 to 2.01).

## Discussion

In this community-based cohort of UKBB participants, including 26,210 with PCR-confirmed ambulatory Covid-19 cases and 381,063 matched contemporary non-infected controls, we found SARS-CoV-2 infection was associated with an approximately 20-fold increase in VTE risk within 30 days of a positive test. However, this risk was largely attenuated in the fully vaccinated participants who then suffered a breakthrough infection.

Known clinical determinants of VTE, including older age, male sex, and obesity, applied to post-Covid-19 VTE. Complete vaccination was associated with an almost 90% relative risk reduction of Covid-19-related VTE. As expected, vaccination did not impact VTE risk in the uninfected peers. Finally, inherited thrombophilia carriers had an additional double risk of post-Covid-19 VTE compared to non-carriers, equivalent to the excess risk attributable to an increase of 10 years of age.

Our finding of a substantially higher incidence of VTE following ambulatory Covid-19 disagreed with a previous meta-analysis of seven heterogeneous small Covid-19 cohorts, which had suggested that mild Covid-19 was not a risk factor for VTE ^21^. However, our data are in line with larger self-controlled case series studies^22,23^ that better accounted for within-person confounding and consistently showed orders of magnitude increases in VTE risk after SARS-CoV-2 infection.

Public interest and concerns have been placed predominantly on vaccine-related rare thromboembolic events ^24^, which has led to vaccine hesitancy and restrictions on its use.^25^ Our study found that the vaccination can offset SARS-CoV-2-induced VTE risk even if people get a breakthrough infection. This evident benefit should not be ignored in the ongoing global vaccination campaigns.

For the first time, we showed that inherited thrombophilia conferred a double risk of Covid-19-related VTE, echoing clinical findings of elevated factor V activity in severe Covid-19 patients.^26–28^ Although a relatively high proportion of congenital thrombophilia was previously detected in a small pilot study of 87 Covid-19 patients, the minimal sample size precluded any robust inference.^29^

We analyzed rich linked data combining extensive community SARS-CoV-2 testing, well-recorded vaccination status, ambulatory and hospital-based clinical outcomes, as well as large scale genotyping data readily available for UKBB participants. The results of this analysis have many noteworthy implications. First, VTE risk management needs re-evaluation for milder ambulatory Covid-19 patients. With emerging evidence and guidelines focusing on VTE prophylaxis for hospitalized Covid-19 patients, further work is now necessary to mitigate risk in the community. Second, although the aetiology of post-Covid VTE is complex and multifaceted, our findings elucidated the role of Factor V and possibly Prothrombin proteins as contributing factors. Third, although genetic testing of inherited thrombophilia for VTE prevention has been previously discussed in many clinical scenarios,^30,31^ this newly identified effect on Covid-19-related VTE, comparable to a 10-year ageing effect, supports the potential value of targeted genetic screening for thrombophilia in the infected older adults. Fourth, traditional known clinical risk factors of VTE are still informative to identify subgroups of people at high risk of Covid-19-related VTE. Finally, our data demonstrate the significant impact of vaccination to minimize the risk of Covid-19 VTE.

This study also has some limitations. Residual confounding cannot be ruled out in this observational study, although robust statistical approaches for causal inference was applied, including PS matching and knowledge-driven negative control analyses. Participants recruited in UKBB might not fully represent the general population and were likely healthier. Although those Covid-19 participants were from non-hospital settings, they were likely to be symptomatic and more severe cases. The extent to which asymptomatic infection is associated with VTE risk warrants further investigation. Finally, the estimates from our analyses were an average and mixed effect of several SARS-CoV-2 strains, which should be cautiously extrapolated to novel variants, such as Omicron.

Community-based ambulatory Covid-19 is associated with a striking 20-fold excess risk of VTE. This risk was much higher among unvaccinated individuals, and increased with older age, in men, and in patients with obesity. Inherited thrombophilia further doubled VTE risk, comparable to a 10-year ageing effect. Our findings call for targeted prevention strategies for post-Covid VTE in outpatient settings, and suggest an aetiological role of inherited thrombophilia. The clinical utility of genetic testing to stratify infected patients and inform personalized thromboprophylaxis warrants further research.

## Data Availability

Bonafide researchers can apply to use the UK Biobank dataset by registering and applying at http://ukbiobank.ac.uk/register-apply/. The datasets generated and/or analysed during the current study are available from the corresponding author on reasonable request

## Declaration of interests

### Ethical approval

All participants provided written informed consent at the UKBB cohort recruitment. This study received ethical approval from UKBB Ethics Advisory Committee (EAC) and was performed under the application of 65397.

### Data sharing

Bonafide researchers can apply to use the UK Biobank dataset by registering and applying at http://ukbiobank.ac.uk/register-apply/.The datasets generated and/or analysed during the current study are available from the corresponding author on reasonable request.

### Transparency declaration

The lead author affirms that this manuscript is an honest, accurate, and transparent account of the study being reported; that no important aspects of the study have been omitted.

### Contributors

D.P.A., J.Q.X., D.G, R.P were responsible for the study design. J.Q.X. did the data analyses, and A.P.U. checked the statistical codes. J.Q.X. and D.P.A. drafted the manuscript, and all co-authors reviewed and approved it for submission.

**Supplementary Results 1:**
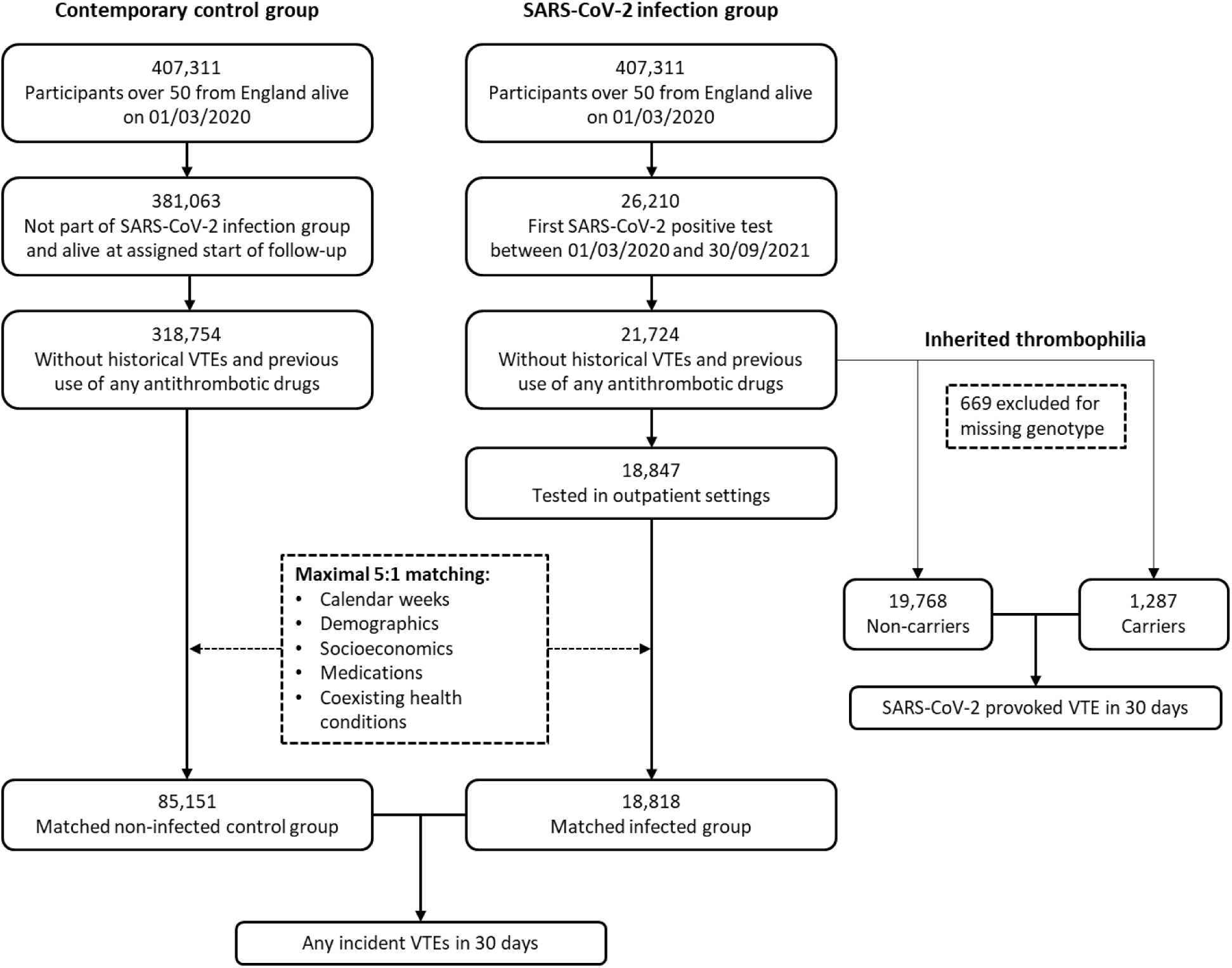
Flow-chart for the study design and participants’ eligibility.

**Supplementary Results 2:**
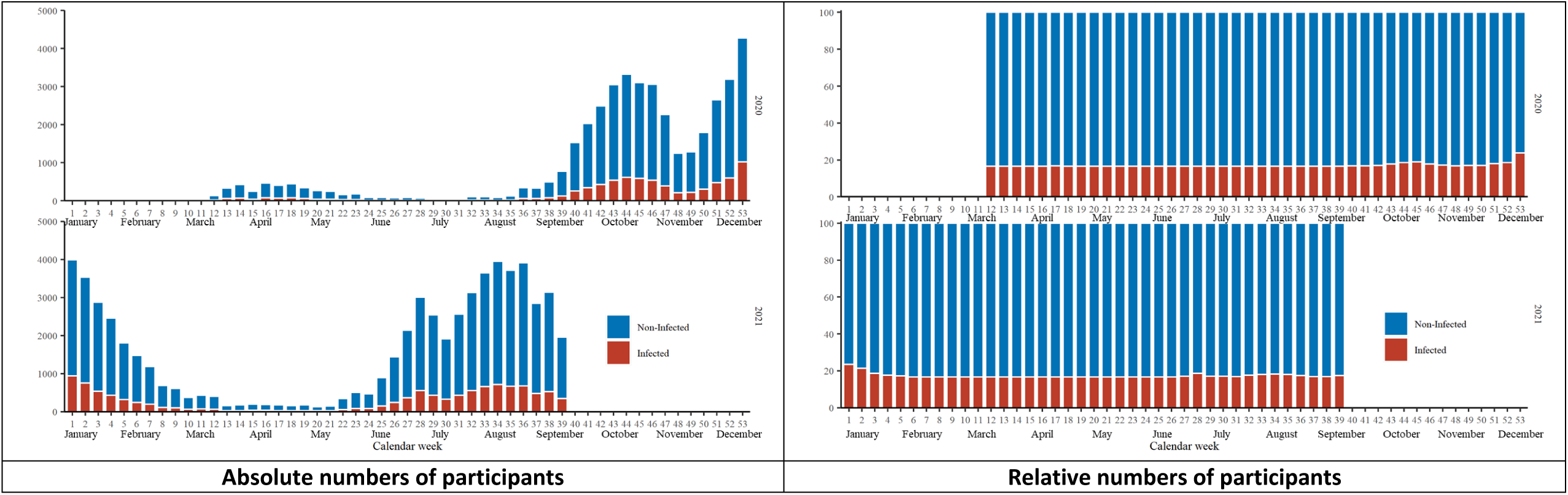
The distribution of infected and 1:5 matched non-infected persons over the study period.

**Supplementary Results 3:**
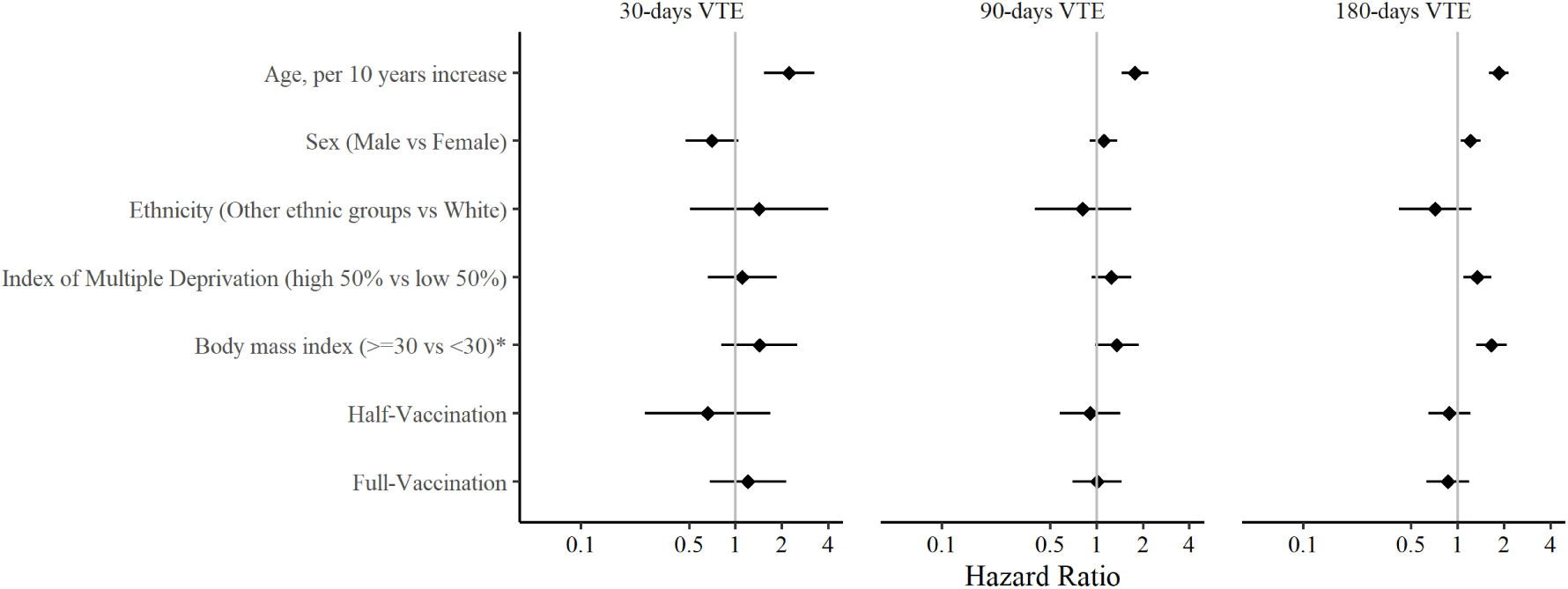
Associations of clinical factors and vaccination status with Covid-19-unrelated VTE (Any VTE events in the non-infected cohort). * Body mass index was measured at cohort recruitment. Because the VTE events in the non-infected group were very rare, we extended follow-up to 180 days to increase statistical power.

**Supplementary Results 4:** The frequency of study genetic variations.

|  | Inherited thrombophilia (any of the two risk variants) | Factor V Leiden risk variant | Prothrombin G20210A risk variant |
| --- | --- | --- | --- |
| All UKBB participants (407,311) | 6.26% | 4.43% | 2.03% |
| Infected participants (21,724) | 6.11% | 4.32% | 1.86% |

**Supplementary Results 5:** Demographic and clinical characteristics of SARS-CoV-2 infected participants, stratified by the inherited thrombophilia.

|  | **Infected participants** |  | **SMD** |
| --- | --- | --- | --- |
|  | **Without inherited thrombophilia** | **With inherited thrombophilia** |  |
| Number | 19,768 | 1,287 |  |
| Age, mean (sd) | 64.82 (8.25) | 64.82 (8.32) | <0.001 |
| Sex, male (%) | 8804 (44.5) | 555 (43.1) | 0.028 |
| Ethnicity White (%) | 16560 (83.8) | 1131 (87.9) | 0.118 |
| Index of Multiple Deprivation, mean (sd) | 19.97 (14.49) | 19.48 (14.71) | 0.034 |
| Body mass index, mean (sd) | 27.78 (4.88) | 27.91 (5.03) | 0.026 |
| **Vaccination status** |  |  |  |
| Not vaccinated | 12426 (62.9) | 802 (62.3) | 0.011 |
| Partial-vaccinated | 956 ( 4.8) | 54 ( 4.2) | 0.031 |
| Fully-vaccinated | 6386 (32.3) | 431 (33.5) | 0.025 |
| **Medications (%)** |  |  |  |
| **(Past one year)** |  |  |  |
| Lipid lowering drugs | 4744 (24.0) | 316 (24.6) | 0.013 |
| RAS inhibitors | 3383 (17.1) | 232 (18.0) | 0.024 |
| Other anti-hypertensives | 1514 ( 7.7) | 92 ( 7.1) | 0.019 |
| Proton pump inhibitors | 5376 (27.2) | 341 (26.5) | 0.016 |
| Diabetes medicines | 1210 ( 6.1) | 70 ( 5.4) | 0.029 |
| Antidepressants | 3278 (16.6) | 228 (17.7) | 0.03 |
| Systemic glucocorticoids | 1102 ( 5.6) | 69 ( 5.4) | 0.009 |
| Immunosuppressants | 220 ( 1.1) | 16 ( 1.2) | 0.012 |
| Antineoplastic agents | 19 ( 0.1) | 0 ( 0.0) | 0.044 |
| **Coexisting conditions (%)** |  |  |  |
| **(Past years)** |  |  |  |
| Cancer | 1576 ( 8.0) | 105 ( 8.2) | 0.007 |
| Malignant cancer | 92 ( 0.5) | 3 ( 0.2) | 0.039 |
| Diabetes (uncomplicated) | 1614 ( 8.2) | 97 ( 7.5) | 0.023 |
| Diabetes (end-organ damage) | 455 ( 2.3) | 29 ( 2.3) | 0.003 |
| Congestive heart failure | 115 ( 0.6) | 8 ( 0.6) | 0.005 |
| Myocardial infarction | 52 ( 0.3) | 2 ( 0.2) | 0.024 |
| Cerebrovascular disease | 164 ( 0.8) | 8 ( 0.6) | 0.025 |
| Peripheral vascular disease | 65 ( 0.3) | 2 ( 0.2) | 0.035 |
| Liver disease (mild) | 93 ( 0.5) | 7 ( 0.5) | 0.01 |
| Liver disease (moderate to severe) | 54 ( 0.3) | 3 ( 0.2) | 0.008 |
| COPD | 3363 (17.0) | 206 (16.0) | 0.027 |
| Chronic kidney disease | 738 ( 3.7) | 58 ( 4.5) | 0.039 |
| Peptic ulcer | 431 ( 2.2) | 25 ( 1.9) | 0.017 |
| Rheumatoid arthritis | 541 ( 2.7) | 32 ( 2.5) | 0.016 |
| Dementia | 254 ( 1.3) | 20 ( 1.6) | 0.023 |
| Hemiplegia | 15 ( 0.1) | 0 ( 0.0) | 0.039 |
| AIDS | 19 ( 0.1) | 0 ( 0.0) | 0.044 |
| **Hospital admissions (numbers, past one year)** | 0.45 (2.33) | 0.39 (1.36) | 0.032 |

**Supplementary Results 6:** Effects of inherited thrombophilia on venous thromboembolism by community and hospitalized Covid-19 cases.

| **Exposure** | **Adjusted HR**<br>**(95% CI)** | **P value** |
| --- | --- | --- |
| **All Covid-19 cases** |  |  |
| Inherited thrombophilia | 2.05 (1.15 to 3.66) | 0.01* |
| Factor V Leiden | 2.17 (1.13 to 4.15) | 0.02* |
| Prothrombin G20210A | 1.52 (0.48 to 4.79) | 0.45 |
| **Community Covid-19 cases** |  |  |
| Inherited thrombophilia | 1.60 (0.69 to 3.72) | 0.27 |
| Factor V Leiden | 1.89 (0.76 to 4.72) | 0.17 |
| Prothrombin G20210A | 0.83 (0.12 to 6.01) | 0.855 |
| **Hospital Covid-19 cases** |  |  |
| Inherited thrombophilia | 2.82 (1.26 to 6.30) | 0.01* |
| Factor V Leiden | 2.49 (0.98 to 6.29) | 0.05 |
| Prothrombin G20210A | 3.18 (0.77 to 13.15) | 0.10 |

**Supplementary Appendix:** Summary statistics for 297 candidate SNPs used for constructing the polygenic risk score for VTE.

| Chr:Pos | rsid | Effect Allele | Beta |
| --- | --- | --- | --- |
| 1:11910677 | rs632793 | G | 0.0466 |
| 1:150388318 | rs698915 | A | 0.0561 |
| 1:166533517 | rs764024 | T | 0.0502 |
| 1:168406710 | rs111414961 | A | 0.1183 |
| 1:168500034 | rs72703796 | G | 0.2014 |
| 1:168572720 | rs72705895 | T | 0.1313 |
| 1:168577239 | rs12567872 | T | 0.0893 |
| 1:168619548 | rs1933116 | T | 0.1724 |
| 1:168673803 | rs611769 | C | 0.0562 |
| 1:168714137 | rs10918970 | T | 0.0572 |
| 1:168729611 | rs12117978 | A | 0.2166 |
| 1:168812398 | rs34258243 | G | 0.1015 |
| 1:168889373 | rs78516619 | G | 0.6841 |
| 1:168909952 | rs7526462 | T | 0.0714 |
| 1:168937072 | rs113123846 | G | 0.4417 |
| 1:168960286 | rs1322487 | A | 0.046 |
| 1:168969254 | rs35990973 | A | 0.1057 |
| 1:169012124 | rs1320969 | G | 0.0478 |
| 1:169014610 | rs116140155 | A | 0.2498 |
| 1:169031755 | rs113079063 | T | 0.2356 |
| 1:169038856 | rs12747018 | T | 0.0717 |
| 1:169064630 | rs1200118 | G | 0.1111 |
| 1:169070213 | rs1892091 | C | 0.0526 |
| 1:169079419 | rs1358714 | A | 0.0444 |
| 1:169086013 | rs2143289 | T | 0.2335 |
| 1:169107377 | rs7520186 | T | 0.0873 |
| 1:169276816 | rs10732287 | T | 0.0916 |
| 1:169316610 | rs10158131 | G | 0.0673 |
| 1:169318242 | rs72702145 | A | 0.1342 |
| 1:169463296 | rs147474835 | G | 0.12 |
| 1:169463519 | rs4656683 | T | 0.1781 |
| 1:169486141 | rs966751 | G | 0.1142 |
| 1:169511555 | rs6032 | T | 0.1351 |
| 1:169593113 | rs3917862 | G | 0.2198 |
| 1:169620673 | rs12075684 | G | 0.0534 |
| 1:169659128 | rs185120584 | T | 0.5696 |
| 1:170051841 | rs115476742 | C | 0.0857 |
| 1:170080014 | rs12128208 | C | 0.1373 |
| 1:170174298 | rs12144655 | A | 0.0893 |
| 1:181031783 | rs3936939 | A | 0.0458 |
| 1:201882277 | rs2644120 | C | 0.058 |
| 1:207282149 | rs2842700 | A | 0.1138 |
| 1:207285043 | rs577695638 | C | 0.1708 |
| 1:230417394 | rs3088075 | T | 0.0798 |
| 1:248028780 | rs79322592 | A | 0.0517 |
| 1:248039451 | rs3811444 | C | 0.0513 |
| 1:9341786 | rs677665 | T | 0.0521 |
| 2:127934102 | rs10198483 | A | 0.0559 |
| 2:127979335 | rs111736896 | A | 0.0717 |
| 2:128133076 | rs35293119 | G | 0.0442 |
| 2:128175875 | rs1799809 | G | 0.072 |
| 2:128187428 | rs67495946 | C | 0.0631 |
| 2:128398892 | rs67076363 | A | 0.0733 |
| 2:161845453 | rs75145714 | C | 0.1264 |
| 2:178238316 | rs4893934 | G | 0.0449 |
| 2:198894550 | rs535527575 | G | 0.1319 |
| 2:198946551 | rs6434955 | G | 0.0587 |
| 2:68530180 | rs13013670 | T | 0.0485 |
| 2:68619981 | rs1867312 | C | 0.0585 |
| 3:126238371 | rs11721316 | A | 0.061 |
| 3:126276710 | rs12636448 | C | 0.0456 |
| 3:150872174 | rs75347181 | A | 0.1323 |
| 3:150880908 | rs3116549 | C | 0.0522 |
| 3:185234111 | rs115110838 | T | 0.0649 |
| 3:194787481 | rs116764841 | T | 0.0661 |
| 3:36140951 | rs113114306 | T | 0.1047 |
| 3:39240559 | rs7622284 | T | 0.0741 |
| 3:6084766 | rs11712865 | G | 0.0799 |
| 3:77048575 | rs6799348 | G | 0.1488 |
| 3:88354842 | rs1841009 | T | 0.0567 |
| 3:89250573 | rs7374904 | A | 0.0751 |
| 3:89679389 | rs141912156 | T | 0.1656 |
| 3:90390344 | rs9858006 | A | 0.0465 |
| 3:93518545 | rs139156297 | T | 0.1588 |
| 3:93580976 | rs9290378 | T | 0.0509 |
| 3:94209055 | rs9290227 | T | 0.0849 |
| 4:155251071 | rs116333064 | A | 0.1157 |
| 4:155424231 | rs4323084 | T | 0.1012 |
| 4:155432944 | rs4547780 | G | 0.064 |
| 4:155450158 | rs13435192 | T | 0.1124 |
| 4:155474683 | rs115999709 | G | 0.1456 |
| 4:155494926 | rs79726896 | G | 0.1598 |
| 4:155513276 | rs2070008 | T | 0.0975 |
| 4:155515486 | rs1984906 | G | 0.0671 |
| 4:155538470 | rs13130318 | G | 0.1984 |
| 4:155542926 | rs59234924 | G | 0.1077 |
| 4:155544958 | rs72681247 | T | 0.2163 |
| 4:155553541 | rs79201245 | C | 0.1462 |
| 4:187114479 | rs10866290 | C | 0.0464 |
| 4:187130620 | rs72646294 | A | 0.1947 |
| 4:187136519 | rs113435394 | C | 0.2091 |
| 4:187161283 | rs551251712 | C | 0.2385 |
| 4:187169469 | rs2203111 | G | 0.0901 |
| 4:187178014 | rs3775303 | T | 0.1486 |
| 4:187180115 | rs4253333 | G | 0.0494 |
| 4:187190285 | rs925452 | A | 0.2681 |
| 4:187196853 | rs4253414 | C | 0.1544 |
| 4:187200550 | rs56810541 | T | 0.1975 |
| 4:187205929 | rs4253425 | C | 0.2399 |
| 4:187213360 | rs72712610 | A | 0.1509 |
| 4:187219599 | rs56379917 | A | 0.1638 |
| 4:187220894 | rs13126546 | T | 0.1142 |
| 4:187234735 | rs72712626 | G | 0.1365 |
| 4:187240280 | rs13137269 | T | 0.0617 |
| 4:187267791 | rs75440104 | C | 0.1177 |
| 4:187347217 | rs7685922 | C | 0.054 |
| 4:79917638 | rs140303646 | T | 0.218 |
| 5:143119511 | rs325247 | C | 0.0439 |
| 5:172785895 | rs12234072 | G | 0.0661 |
| 5:32831939 | rs12656497 | T | 0.0438 |
| 5:38708554 | rs16867574 | C | 0.059 |
| 5:63916834 | rs4700642 | A | 0.0491 |
| 5:75992254 | rs56347914 | C | 0.0843 |
| 6:147701133 | rs9373523 | G | 0.06 |
| 6:169633335 | rs11759438 | C | 0.0436 |
| 6:25531133 | rs214057 | C | 0.0446 |
| 6:28436060 | rs2531815 | T | 0.047 |
| 6:29894392 | rs1627764 | G | 0.0525 |
| 6:30639412 | rs3094094 | A | 0.0793 |
| 6:31092767 | rs3095304 | T | 0.0634 |
| 6:31239869 | rs2074492 | T | 0.051 |
| 7:151028181 | rs11981586 | C | 0.0657 |
| 7:157763424 | rs1347390 | G | 0.0515 |
| 8:102881195 | rs118105926 | A | 0.1379 |
| 8:106573309 | rs7341574 | T | 0.0452 |
| 8:106590705 | rs4541868 | C | 0.0876 |
| 8:108291878 | rs4236786 | C | 0.0512 |
| 8:108340982 | rs7004172 | G | 0.051 |
| 8:108347806 | rs6991048 | T | 0.0904 |
| 8:27810577 | rs2685413 | G | 0.0608 |
| 8:27820792 | rs10087301 | A | 0.0632 |
| 8:30265541 | rs117564659 | G | 0.1314 |
| 8:53204323 | rs138757339 | C | 0.2102 |
| 8:78572803 | rs17383689 | G | 0.0833 |
| 8:87143573 | rs7812868 | C | 0.0621 |
| 9:116253293 | rs189064188 | T | 0.0804 |
| 9:124362398 | rs146383320 | T | 0.1278 |
| 9:135985796 | rs3761824 | C | 0.0538 |
| 9:136025460 | rs3888561 | C | 0.0559 |
| 9:136031918 | rs7027827 | A | 0.0729 |
| 9:136056956 | rs10793953 | G | 0.0496 |
| 9:136062437 | rs10441806 | C | 0.105 |
| 9:136069931 | rs7039497 | G | 0.1081 |
| 9:136077004 | rs11244032 | C | 0.0837 |
| 9:136080512 | rs149189328 | C | 0.2669 |
| 9:136081319 | rs11244035 | T | 0.2223 |
| 9:136098498 | rs28470788 | T | 0.0723 |
| 9:136121303 | rs7855466 | T | 0.1054 |
| 9:136124590 | rs78755596 | A | 0.2855 |
| 9:136128731 | rs11244051 | A | 0.384 |
| 9:136144593 | rs66697526 | G | 0.1399 |
| 9:136145404 | rs9411377 | A | 0.308 |
| 9:136152070 | rs8176634 | G | 0.1117 |
| 9:136152722 | rs8176630 | C | 0.1382 |
| 9:136156064 | rs55988407 | G | 0.1664 |
| 9:136156230 | rs78590974 | T | 0.2221 |
| 9:136157037 | rs557317 | A | 0.1499 |
| 9:136177993 | rs4962043 | G | 0.0607 |
| 9:136184782 | rs9411395 | G | 0.1408 |
| 9:136184985 | rs11791119 | T | 0.1052 |
| 9:136185324 | rs11789139 | G | 0.1669 |
| 9:136193356 | rs76771223 | G | 0.1431 |
| 9:136212168 | rs117119759 | A | 0.2099 |
| 9:136226421 | rs141397052 | G | 0.2259 |
| 9:136240304 | rs3124755 | C | 0.1278 |
| 9:136255149 | rs62575992 | C | 0.0967 |
| 9:136268084 | rs3124747 | A | 0.0757 |
| 9:136270538 | rs41302673 | G | 0.242 |
| 9:136277854 | rs2285488 | G | 0.1007 |
| 9:136296530 | rs149181677 | T | 0.2461 |
| 9:136311017 | rs652600 | A | 0.0992 |
| 9:136323826 | rs3094373 | A | 0.2432 |
| 9:136343647 | rs3094326 | G | 0.1027 |
| 9:136359182 | rs736418 | G | 0.0556 |
| 9:136365146 | rs28615587 | T | 0.078 |
| 9:136382716 | rs9802874 | A | 0.0636 |
| 9:136390015 | rs13300181 | A | 0.0616 |
| 9:136397195 | rs11507716 | T | 0.0802 |
| 9:136509514 | rs1611128 | G | 0.0495 |
| 10:121010256 | rs10886430 | G | 0.1289 |
| 10:32397591 | rs211416 | T | 0.0684 |
| 10:45632668 | rs2211163 | A | 0.0724 |
| 10:71144324 | rs2305196 | A | 0.0555 |
| 10:71148728 | rs12416320 | G | 0.1553 |
| 10:71153882 | rs3793846 | T | 0.0572 |
| 10:71181371 | rs36054387 | C | 0.0729 |
| 10:71215107 | rs137936874 | G | 0.2231 |
| 10:71218059 | rs10998791 | A | 0.0677 |
| 10:71245276 | rs78707713 | T | 0.2414 |
| 10:71262048 | rs1665581 | G | 0.0771 |
| 10:71333897 | rs12785008 | C | 0.0691 |
| 10:71346272 | rs12783163 | A | 0.0596 |
| 10:76189250 | rs140438685 | A | 0.2024 |
| 10:80898969 | rs1769758 | G | 0.0436 |
| 10:96011865 | rs1547643 | G | 0.0476 |
| 11:126300537 | rs11600151 | T | 0.0793 |
| 11:32967270 | rs563259534 | T | 0.1048 |
| 11:33247621 | rs2061997 | T | 0.049 |
| 11:46559730 | rs11038913 | T | 0.1126 |
| 11:46745003 | rs5896 | T | 0.0668 |
| 11:46893108 | rs2306029 | T | 0.0641 |
| 11:46896126 | rs72897640 | T | 0.0672 |
| 11:47373425 | rs2856656 | C | 0.3976 |
| 11:47794348 | rs34953939 | A | 0.0931 |
| 11:48865680 | rs369876615 | C | 0.1184 |
| 11:50242788 | rs117653193 | T | 0.1049 |
| 11:51282525 | rs11606922 | C | 0.1115 |
| 11:55436134 | rs72910502 | C | 0.1161 |
| 11:56526894 | rs543926510 | C | 0.1339 |
| 11:56735815 | rs75348906 | A | 0.0695 |
| 11:56875074 | rs141798115 | T | 0.2207 |
| 11:61489705 | rs198428 | A | 0.0464 |
| 11:61571348 | rs174548 | C | 0.0842 |
| 11:61621611 | rs73487492 | A | 0.1034 |
| 11:73283937 | rs12274057 | T | 0.0751 |
| 12:104147207 | rs3751198 | G | 0.0491 |
| 12:111932800 | rs7137828 | C | 0.0424 |
| 12:123861452 | rs28413626 | G | 0.0739 |
| 12:32844798 | rs61926202 | A | 0.2109 |
| 12:39156743 | rs137870902 | T | 0.2329 |
| 12:54734289 | rs11170877 | A | 0.0696 |
| 12:6071943 | rs139727584 | C | 0.1076 |
| 12:6150824 | rs183356 | A | 0.1405 |
| 12:6160614 | rs7135039 | T | 0.0781 |
| 12:6170645 | rs78915411 | G | 0.0823 |
| 13:113787459 | rs3211752 | G | 0.06 |
| 13:113808274 | rs12858483 | G | 0.0588 |
| 14:100108918 | rs12886724 | G | 0.0501 |
| 14:26690604 | rs77398404 | T | 0.0794 |
| 14:66082793 | rs2229678 | C | 0.1842 |
| 14:83281618 | rs112089121 | G | 0.0752 |
| 14:92217670 | rs61988257 | A | 0.0603 |
| 14:94838142 | rs112635299 | T | 0.1463 |
| 15:43757184 | rs190543502 | T | 0.1973 |
| 15:43911751 | rs115384559 | T | 0.3853 |
| 15:44880783 | rs148770227 | C | 0.1837 |
| 15:65114833 | rs35204896 | G | 0.0738 |
| 15:66430422 | rs74245462 | T | 0.0503 |
| 15:96125226 | rs17502085 | A | 0.0525 |
| 16:1405044 | rs116468525 | A | 0.2059 |
| 16:81840709 | rs34603417 | A | 0.0456 |
| 16:81844607 | rs9937779 | C | 0.0568 |
| 16:81870969 | rs12445050 | T | 0.1248 |
| 16:81874200 | rs61374069 | A | 0.0524 |
| 16:81896523 | rs55909816 | C | 0.0437 |
| 16:81902990 | rs4889419 | G | 0.0575 |
| 16:81915832 | rs11150422 | G | 0.053 |
| 16:81971403 | rs1071644 | C | 0.0475 |
| 16:81976177 | rs16956040 | C | 0.0613 |
| 16:89265466 | rs12926888 | A | 0.0449 |
| 17:1966457 | rs1048483 | T | 0.0616 |
| 17:2172753 | rs216181 | A | 0.072 |
| 17:43758898 | rs57222984 | G | 0.0522 |
| 17:67081278 | rs77542162 | G | 0.1932 |
| 17:7785590 | rs78209469 | T | 0.0867 |
| 17:8393900 | rs2270744 | C | 0.0447 |
| 18:74465347 | rs62112094 | A | 0.059 |
| 18:75283432 | rs2032276 | G | 0.0435 |
| 18:8800723 | rs631126 | C | 0.0579 |
| 19:10639312 | rs8100818 | T | 0.0483 |
| 19:10659971 | rs11668544 | G | 0.1314 |
| 19:10688153 | rs187758170 | A | 0.1465 |
| 19:10734951 | rs8110479 | C | 0.1131 |
| 19:10741622 | rs12710257 | G | 0.1164 |
| 19:10898413 | rs8107372 | T | 0.0515 |
| 19:17004049 | rs1054533 | T | 0.0451 |
| 19:33896432 | rs4805881 | C | 0.0544 |
| 19:3464793 | rs142170418 | C | 0.1574 |
| 19:45426792 | rs141622900 | A | 0.1007 |
| 19:46268902 | rs2341097 | T | 0.0502 |
| 19:49241006 | rs12981072 | C | 0.0469 |
| 19:55536595 | rs1613662 | A | 0.0818 |
| 19:7832001 | rs874492 | A | 0.0481 |
| 20:22672552 | rs6137727 | A | 0.0534 |
| 20:22938940 | rs62204096 | A | 0.1565 |
| 20:23000653 | rs6076004 | T | 0.0472 |
| 20:23077117 | rs149439892 | A | 0.1527 |
| 20:23170450 | rs34397775 | A | 0.084 |
| 20:23182559 | rs6083037 | A | 0.0798 |
| 20:32485961 | rs6087538 | C | 0.0585 |
| 20:32523172 | rs6059574 | G | 0.0479 |
| 20:33434252 | rs56244533 | A | 0.0567 |
| 20:33435161 | rs17092148 | G | 0.0612 |
| 20:33451060 | rs75627267 | T | 0.1209 |
| 20:33587569 | rs551986443 | C | 0.1488 |
| 20:33612647 | rs564783003 | C | 0.1488 |
| 20:33733180 | rs2050652 | G | 0.0564 |
| 20:33762035 | rs2069946 | C | 0.1255 |
| 20:33772243 | rs6060288 | A | 0.1178 |
| 20:33858772 | rs6058218 | G | 0.0552 |
| 20:33895947 | rs6058227 | T | 0.0813 |
| 20:34025983 | rs143383 | G | 0.0482 |
| 20:34562935 | rs185663249 | A | 0.1487 |
| 20:34712310 | rs6141600 | C | 0.0482 |
| 22:42461918 | rs2854827 | G | 0.0589 |
| 22:43107837 | rs9620086 | G | 0.0609 |
| 22:44324730 | rs738408 | C | 0.0566 |
| 1:169519049 | rs6025 | T | 0.4031 |
| 11:46761055 | rs1799963 | A | 0.2741 |

